# Trends in underlying causes of death in solid organ transplant recipients between 2010 and 2020: Using the CLASS method for determining specific causes of death

**DOI:** 10.1101/2022.01.17.22269434

**Authors:** Andreas Søborg, Joanne Reekie, Allan Rasmussen, Caspar Da Cunha-Bang, Finn Gustafsson, Kasper Rossing, Michael Perch, Paul S. Krohn, Søren S. Sørensen, Thomas K. Lund, Vibeke R. Sørensen, Christina Ekenberg, Louise Lundgren, Isabelle P. Lodding, Kasper S. Moestrup, Jens D. Lundgren, Neval E. Wareham

## Abstract

Monitoring specific underlying causes of death in solid organ transplant (SOT) recipients is important in order to identify emerging trends and health challenges.

This retrospective cohort study includes all SOT recipients transplanted at Rigshospitalet between January 1st, 2010 and December 31st, 2019. The underlying cause of death was determined using the newly developed Classification of Death Causes after Transplantation (CLASS) method. Cox regression analyses assessed risk factors for all-cause and cause-specific mortality.

Of the 1774 SOT recipients included, 299 patients died during a total of 7511 person-years of follow-up (PYFU) with cancer (N=57, 19%), graft rejection (N=55, 18%) and infections (N=52, 17%) being the most frequent causes of death.

We observed a lower risk of all-cause death with increasing transplant year (HR 0.91, 95% CI 0.86-0.96 per 1-year increase), alongside death from graft rejection (HR 0.84 per year, 95% CI 0.74-0.95) and death from infections (HR 0.87 per year, 95% CI 0.77-0.98).

Further, there was a trend towards lower cumulative incidence of death from cardiovascular disease, graft failure and cancer in more recent years, while death from other organ specific and non-organ specific causes did not decrease.

All-cause mortality among SOT recipients has decreased over the past decade, mainly due to a decrease in graft rejection- and infection-related deaths. Conversely, causes from a broad range of diseases have remained unchanged, suggesting that cause of death among SOT recipients is increasingly diverse and warrants a multidisciplinary effort and attention in the future.

## Introduction

Survival after solid organ transplantation (SOT) has increased since the 1980s and 1990s[1, 2]. The primary improvement has been observed in short term survival within the first year after transplantation, due to a decrease in death from predominately graft failure and a decrease in death from infections across all types of solid organ transplantations[3, 4]. This is likely a result of novel immunosuppressive regimens, improved surgical treatment and implementation of preemptive initiatives towards infections[5-7].

However, overall mortality rates among individuals who have received SOT remains far above those observed in the age-matched general population[8, 9]. Some of this excess risk could be attributed to an increase in pre-transplant comorbidities and risk factors including older age at time of transplant, in recipients transplanted in more recent years[2, 10, 11]. In 2019 for example, 21.8% of kidney transplant recipients in the United States were aged 65 years or older compared to just 9.2% in 2000[12]. As the age of SOT recipients increases, recipients become more vulnerable to multiple diseases and illnesses. For example, deaths due to cancer have become an emerging cause of death in SOT recipients[13-15].

A common limitation in previous studies has been incomplete classification of causes of death with up to 60% of deaths attributed to unknown or missing causes in large cohort studies[13, 16-19]. Further, using national death causes registries where cause of death more often refers to the underlying disease leading to transplantation may be misleading and thus less useful in a transplant setting[20]. Therefore, the aim of this work was to determine the specific underlying causes of death in SOT recipients using the recently developed Classification of Death Causes after Transplantation (CLASS) method[20], which includes a thorough investigation of the events leading to death and a review process by independent transplant specialists. This method has previously shown relatively high discrepancy when compared with national registries, resulting in reclassification of causes of death in more than 60% of cases[20]. A further aim was to investigate changes in both all-cause and specific causes of death observed during the last 10 years in SOT recipients from the largest transplantation center in Denmark.

## Materials and methods

### Study design and population

We conducted a single center, observational cohort study of SOT recipients (heart, kidney, liver, lung or pancreas transplantations) at Rigshospitalet, a large tertiary transplant center in Copenhagen, Denmark with regional function for transplantation of kidneys and adult hearts and national function for transplantation of all other solid organs as well as pediatric hearts. All children and adults who received their first solid organ transplant between January 1^st^, 2010 and December 31^st^, 2019 and enrolled in the Management of Post-Transplant Infections in Collaborating Hospitals (MATCH) program[21, 22] were included. A small number of patients transplanted abroad or at another Danish transplant center and subsequently enrolled in MATCH for monitoring were included as well.

### CLASS method

Patients who died during the 10-year period of follow-up were registered both continuously through reporting by clinicians responsible of care and retrospectively through linkage to the Danish Civil Registration System where data on deaths in Denmark are close to 100% complete[23]. The specific underlying cause of death was obtained in accordance with a modified version of the validated Classification of Death Causes after Transplantation (CLASS) method[20] which includes completion of a Case Record Form for deceased patients and a review process by two independent and mutually anonymous transplant specialists who each review and decide the cause of death chosen from a list of more than 250 specific causes[24]. In case of disagreement between the two reviewers, an adjudication process is initiated where the cause of death can be discussed and agreed upon. Ultimately, if agreement between the two reviewers cannot be achieved, a panel of specialists will determine the final cause of death. The modification consisted of fatal cases being considered either complicated or uncomplicated based on criteria proposed previously[20] and only cases categorized as complicated being sent into review.

### Data sources

Electronic medical files were reviewed upon completion of Case Record Forms. Further, clinical characteristics, biochemical parameters, pathology, microbiology, imaging, and all diagnoses registered during hospital admission were retrieved from the Centre of Excellence for Personalized Medicine of Infectious Complications in Immune Deficiency (PERSIMUNE)[25] data repository. The PERSIMUNE data repository contains data from national and regional registries as well as clinical databases collected prospectively as part of clinical care (S1 Table).

### Definitions

*Underlying cause of death* was defined as the disease or comorbidity leading to the death or directly causing the event classified as the immediate cause of death. Underlying causes of death were further grouped in wider categories to perform statistical analyses, including deaths due to cardiac or vascular disease, graft failure, graft rejection, infections, cancer, unknown causes and other organ specific causes (aside from those specified above) and non-organ specific causes. Non-organ specific causes include hemorrhage, alcohol abuse and other causes. Deaths from cancer include de novo cancer (primary and secondary), progression of existing cancer and relapse of cancer leading to transplantation. Graft-versus-host disease (GvHD), a rare cause of death in SOT recipients, was grouped with graft rejection. In cases where graft rejection led to subsequent graft failure and death of the patient, underlying cause of death was classified as graft rejection.

Patients with pancreas transplantations or simultaneous liver and other organ transplantations were grouped as liver/pancreas transplant recipients as immunosuppressive medication followed the same algorithm as liver recipients. Further, patients with simultaneous lung and kidney transplantations were grouped as lung transplant recipients for the same reason.

The study period was divided into three posttransplant periods for investigation of changes in mortality over time after transplantation, namely an early period (≤1 year posttransplant), an intermediate period (between >1 and 3 years posttransplant) and a late period (between >3 and 10 years posttransplant).

To assess changes in mortality according to transplant year, the study period was divided into an early (2010-14) and a recent (2015-19) transplant era. For changes between transplant eras, only deaths within the first five years posttransplant were assessed due to limited observation period for those transplanted more recently.

### Statistical analysis

Patients were included at their first date of transplantation and followed until date of death or censor at December 31^st^, 2019 (the last date which records were available).

Patient characteristics at the time of transplant were described as frequencies and percentages for categorical variables, and continuous data were reported as medians with interquartile ranges (IQR). Mann–Whitney U testing was used to determine the differences in medians, and the chi-square test was used to test the frequency distributions.

Cumulative incidence curves for cause-specific mortality (wider categories) were calculated for all-types of transplantations and for each of the three posttransplant periods including only patients alive at the beginning of each period. Cumulative incidence curves were also performed for specific causes of death stratified by early (2010-14) and recent (2015-19) transplant era.

Crude mortality rates per 1000 person-years of follow-up (PYFU) for all-cause and cause-specific mortality according to transplanted organ were estimated.

Univariable and multivariable Cox proportional hazard models were conducted to investigate the association between baseline and time-updated characteristics with all-cause and cause-specific mortality, with deaths from other causes treated as competing risks. Baseline characteristics included gender, transplanted organ, age at time of transplant (categorized as age<18, age 18-44, age 45-65 and age>65) and transplant year (per 1-year increase), while number of transplantations (1 or >1) was included as a time-updated variable. Number of transplantations count simultaneous transplantations as one, and only transplantations performed during the study period are counted, as patients with a transplantation prior to 2010 were excluded from the cohort. Due to their clinical relevance, all baseline characteristics were included in the multivariable analyses regardless of their significance in the univariate models. All analyses performed using Cox models were repeated as Fine and Gray models with no deviation of any significance in the results observed between the two types of test. All tests performed were reported with two-tailed p-values.

Sensitivity analyses in a sub-group of patients transplanted before 2018 were conducted to investigate if similar patterns were observed in participants with at least two years of follow-up. In additional sensitivity analyses, pre-transplant chronic diseases were included in multivariable analyses, i.e. cardiovascular disease, diabetes mellitus, chronic lung disease, chronic kidney disease, chronic liver disease, connective tissue disease, cerebrovascular disease, cancer, and peripheral vascular disease. National pre-transplant chronic disease data were available until 2017, hence these sensitivity analyses censored data after 2017.

Categories of causes of death were only assessed if there were more than 30 deaths with that cause.

All data analyses were performed using SAS Enterprise Guide version 7.1 [SAS Institute, Cary, NC] and plots were created using GraphPad Prism 9 [GraphPad Software, San Diego, CA].

### Approvals

Relevant approvals for this study were obtained from the Danish National Data Protection Agency (2012-58-0004, RH-2016-47, with I-Suite number: I-suite 04433), the Danish Patient Safety authority 31-1521-160 and the MATCH steering committee. According to Danish legislations, informed consent is not required for this type of study.

## Results

### Patient population and demographic changes over transplant year

A total of 1774 patients in the MATCH cohort transplanted during the study period were included. Of these, kidneys were the most frequently transplanted organ (N=837), followed by liver/pancreas (N=505), lung (N=299), and heart (N=133) (Table 1). The majority of the patients were males (59.9%) and median age at time of transplantation was 49.6 years (IQR 38.1 – 58.5). In the more recent transplant era (2015-19) there was a relative increase in the number of liver/pancreas transplantations compared to the earlier era (2010-2014) (N=289 vs N=216, p=0.005) and a relative decrease in the number of kidney transplantations (N=425 vs. N=412, p=0.036). The number of patients included in the cohort was similar across the two transplant eras (N=854 and N=920), and no changes between eras were seen in the distribution of gender (p=0.344), age groups (p=0.459), median age (p=0.107) and baseline chronic diseases. Baseline characteristics are described in Table 1.

**Table 1.** Cohort demographics at the time of transplantation (except time-updated number of transplantations).

| Characteristics | All transplants | Heart transplant | Kidney transplant | Liver/pancreas transplant* | Lung transplant** |
| --- | --- | --- | --- | --- | --- |
| Male; N (%) | 1062 (59.9) | 91 (68.4) | 531 (63.4) | 285 (56.4) | 155 (51.8) |
| Female; N (%) | 712 (40.1) | 42 (31.6) | 306 (36.6) | 220 (43.6) | 144 (48.2) |
| Age < 18 years; N (%) | 119 (6.7) | 13 (9.8) | 37 (4.4) | 65 (12.9) | 4 (1.3) |
| Age 18-44 years; N (%) | 540 (30.4) | 38 (28.6) | 268 (32.0) | 154 (30.5) | 80 (26.8) |
| Age 45-65 years; N (%) | 963 (54.3) | 72 (54.1) | 419 (50.1) | 260 (51.5) | 212 (70.9) |
| Age > 65 years; N (%) | 152 (8.6) | 10 (7.5) | 113 (13.5) | 26 (5.2) | 3 (1.0) |
| Transplanted in early era (2010-14); N (%) | 854 (48.1) | 65 (48.9) | 425 (50.8) | 216 (42.8) | 148 (49.5) |
| Transplanted in late era (2015-19); N (%) | 920 (51.9) | 68 (51.1) | 412 (49.2) | 289 (57.2) | 151 (50.5) |
| Number of transplantations= 1***; N (%) | 1739 (98.0) | 133 (100) | 831 (99.3) | 480 (95.1) | 295 (98.7) |
| Number of transplantations > 1***; N (%) | 35 (2.0) | . | 6 (0.7) | 25 (5.0) | 4 (1.3) |
| Total; N (%) | 1774 (100) | 133 (100) | 837 (100) | 505 (100) | 299 (100) |

\* Liver/pancreas transplant includes single or multiple liver transplants (n=459), single pancreas transplants (n=1), combined liver and kidney transplants (n=17) and combined pancreas and kidney transplants (n=28).
\*\* Lung transplant includes single or multiple lung transplants (n=298) and combined lung and kidney transplants (n=1).
\*\*\* Number of transplantations count simultaneous transplantations as one, and only transplantations performed during the study period (2010-2020) are counted.

During a total of 7511 PYFU, 299 patients died (incidence rate of 39.8 per 1000 PYFU, 95% confidence interval (CI) 35.5-44.6) with the highest both absolute and relative numbers seen in lung transplant recipients (104 deaths in 299 patients, incidence rate of 100.3 per 1000 PYFU, 95% CI 82.8-121.6). Clinical characteristics for deceased patients are presented in Table 2.

**Table 2.**
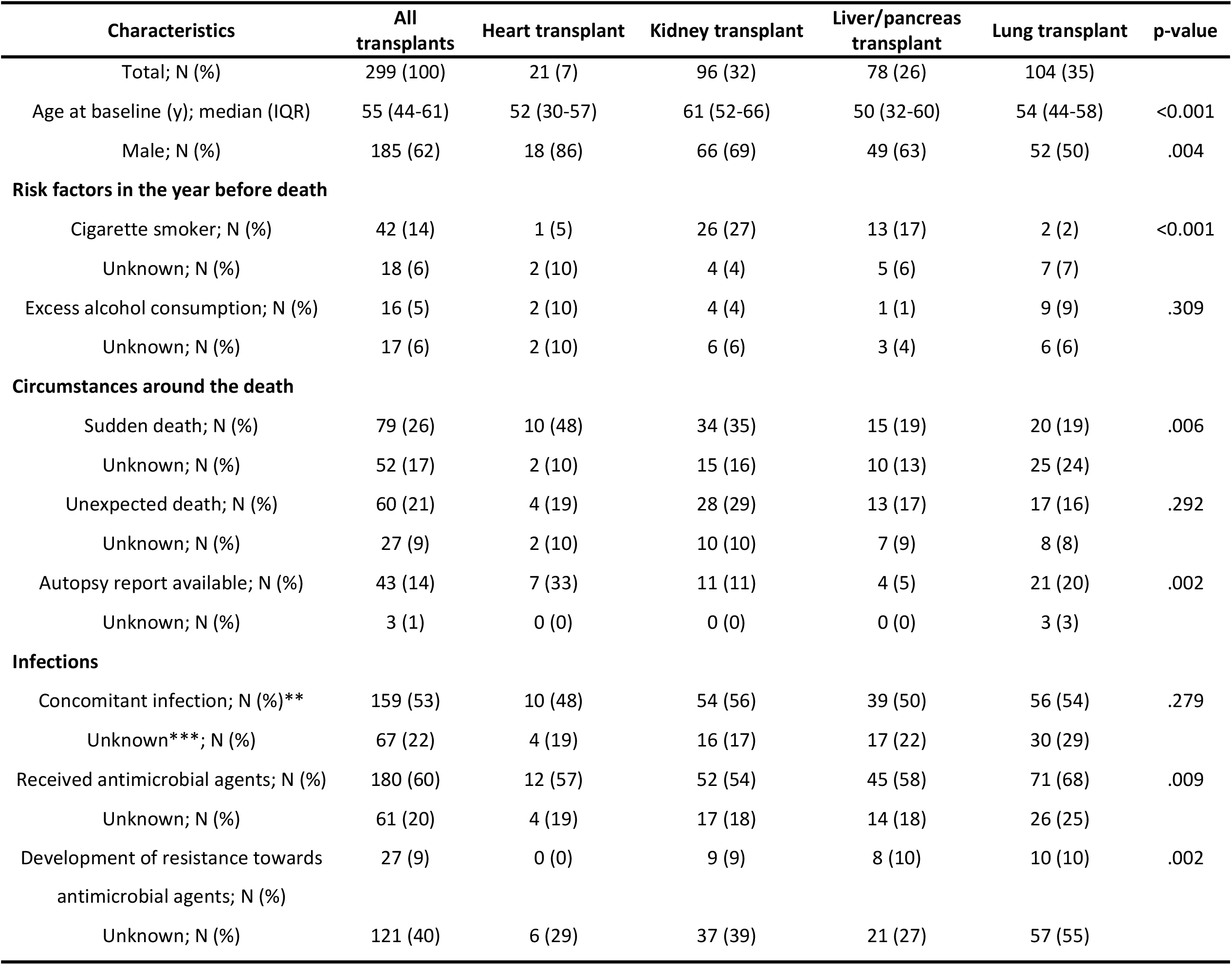
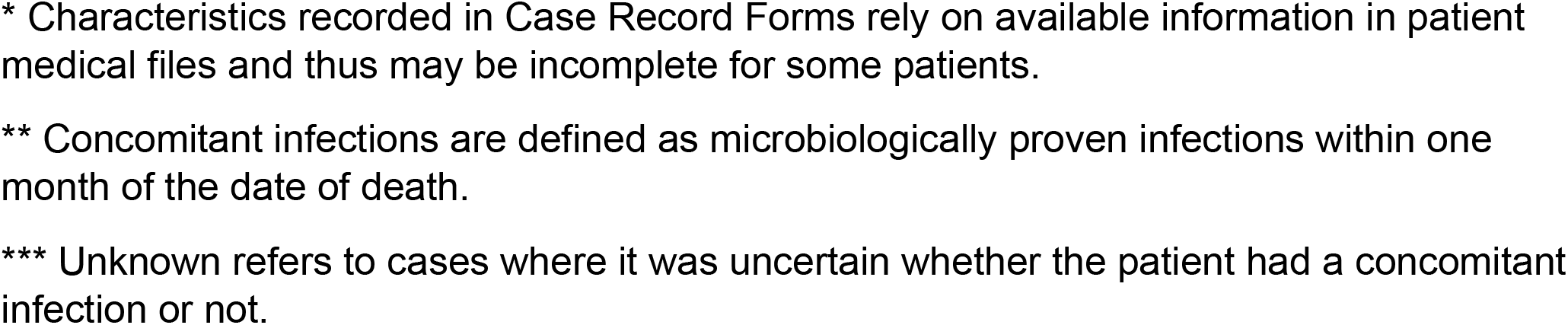
Clinical characteristics of deceased patients recorded in Case Record Forms*.

| Characteristics | All transplants | Heart transplant | Kidney transplant | Liver/pancreas transplant | Lung transplant | p-value |
| --- | --- | --- | --- | --- | --- | --- |
| Total; N (%) | 299 (100) | 21 (7) | 96 (32) | 78 (26) | 104 (35) |  |
| Age at baseline (y); median (IQR) | 55 (44-61) | 52 (30-57) | 61 (52-66) | 50 (32-60) | 54 (44-58) | <0.001 |
| Male; N (%) | 185 (62) | 18 (86) | 66 (69) | 49 (63) | 52 (50) | .004 |
| **Risk factors in the year before death** |  |  |  |  |  |  |
| Cigarette smoker; N (%) | 42 (14) | 1 (5) | 26 (27) | 13 (17) | 2 (2) | <0.001 |
| Unknown; N (%) | 18 (6) | 2 (10) | 4 (4) | 5 (6) | 7 (7) |  |
| Excess alcohol consumption; N (%) | 16 (5) | 2 (10) | 4 (4) | 1 (1) | 9 (9) | .309 |
| Unknown; N (%) | 17 (6) | 2 (10) | 6 (6) | 3 (4) | 6 (6) |  |
| **Circumstances around the death** |  |  |  |  |  |  |
| Sudden death; N (%) | 79 (26) | 10 (48) | 34 (35) | 15 (19) | 20 (19) | .006 |
| Unknown; N (%) | 52 (17) | 2 (10) | 15 (16) | 10 (13) | 25 (24) |  |
| Unexpected death; N (%) | 60 (21) | 4 (19) | 28 (29) | 13 (17) | 17 (16) | .292 |
| Unknown; N (%) | 27 (9) | 2 (10) | 10 (10) | 7 (9) | 8 (8) |  |
| Autopsy report available; N (%) | 43 (14) | 7 (33) | 11 (11) | 4 (5) | 21 (20) | .002 |
| Unknown; N (%) | 3 (1) | 0 (0) | 0 (0) | 0 (0) | 3 (3) |  |
| **Infections** |  |  |  |  |  |  |
| Concomitant infection; N (%)** | 159 (53) | 10 (48) | 54 (56) | 39 (50) | 56 (54) | .279 |
| Unknown***; N (%) | 67 (22) | 4 (19) | 16 (17) | 17 (22) | 30 (29) |  |
| Received antimicrobial agents; N (%) | 180 (60) | 12 (57) | 52 (54) | 45 (58) | 71 (68) | .009 |
| Unknown; N (%) | 61 (20) | 4 (19) | 17 (18) | 14 (18) | 26 (25) |  |
| Development of resistance towards antimicrobial agents; N (%) | 27 (9) | 0 (0) | 9 (9) | 8 (10) | 10 (10) | .002 |
| Unknown; N (%) | 121 (40) | 6 (29) | 37 (39) | 21 (27) | 57 (55) |  |

\* Characteristics recorded in Case Record Forms rely on available information in patient medical files and thus may be incomplete for some patients.
\*\* Concomitant infections are defined as microbiologically proven infections within one month of the date of death.
\*\*\* Unknown refers to cases where it was uncertain whether the patient had a concomitant infection or not.

### Characteristics associated with all-cause mortality and change over transplant year

In multivariable analyses, the risk of all-cause mortality decreased by transplant year (hazard ratio (HR) 0.91, 95% CI 0.86-0.96 per 1-year increase). The decreasing association was also seen in sensitivity models including patients transplanted before 2018 only (HR 0.92, 95% CI 0.87-0.97 per 1-year increase), and in analyses adjusting for pre-transplant chronic diseases (HR 0.92, 95% CI 0.86-0.97 per 1-year increase).

Heart, kidney, and liver/pancreas transplant recipients had significantly lower risk of all-cause mortality compared with lung transplant recipients (S1 Fig). The association between age at time of transplantation and risk of mortality was not linear, with patients aged 65 years or older at baseline having a 2.6-fold higher risk of dying when compared to patients aged <18 years and 45-65 years (HR 2.6, 95% CI 1.5-4.4 and HR 2.6, 95% CI 1.8-3.7, respectively) and a 4.6-fold higher risk compared to those aged 18-44 years (HR 4.6. 95% CI 3.0-7.0). Having more than one transplantation was associated with a 2.1-fold higher risk of mortality (HR 2.1, 95% CI 1.2-3.6), while gender had no significant association with all-cause mortality.

Analyses of the association between baseline characteristics and all-cause mortality stratified by transplanted organ were limited due to small numbers although results were similar to the main analyses. Thus, older patients and having more than one transplantation were associated with higher mortality. However, a lower risk of all-cause mortality by increasing transplant year was observed in lung transplant recipients only (HR 0.86, 95% CI 0.79-0.94 per one-year increase).

### Main causes of death

Cancer (N=57, 19.1%), graft rejection (N=55, 18.4%) and infections (N=52, 17.4%) were the most frequent causes of death overall, with variations between different transplanted organs (Fig 1). In 8.0% of the cases cause of death could not be determined, as the patient died either unobserved outside a hospital or in local hospitals where no hospital records were available for assessment. Late deaths more often had unknown cause of death compared to early deaths.

**Fig 1.**
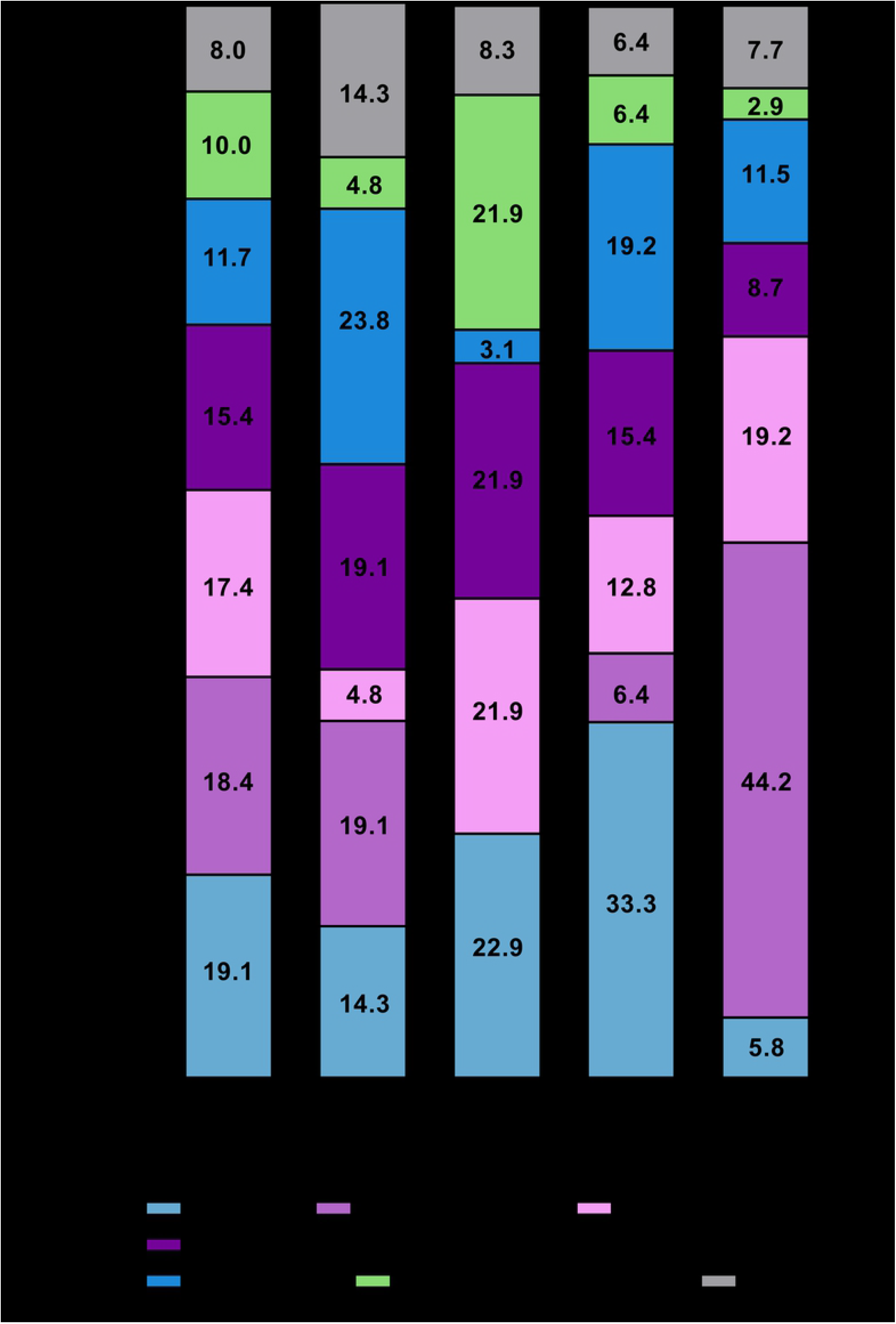
Distribution of recorded causes of death (percentages) between types of transplantation. * Graft rejection includes one death from GvHD. ** Other organ specific and non-organ specific causes includes death from organ failure or dysfunction not caused by graft rejection, graft failure, cancer or infection, death from hemorrhage and death from other causes.

The majority of deaths from graft rejection occurred in lung transplant recipients, where this was the main underlying cause of death with an incidence rate of 44.4 (95% CI 33.2-59.3) per 1000 PYFU (N=46 in 104 deaths). Of these, the main type was chronic rejection (N=39, 84,8%), followed by acute rejection (N=3, 6.5%), unspecified rejection (N=3, 6.5%) and gastrointestinal graft versus host disease (N=1, 2.2%).

Infections were the third most frequent cause of death across all types of transplantations, with an incidence rate of 6.9 (95% CI 5.3-9.1) per 1000 PYFU for the entire cohort. Of the 52 deaths due to infections, 15 (28.8%) were caused by bacterial infection, 9 (17.3%) by viral infection, 7 (13.5%) by fungal infection and 1 (1.9%) by a parasitical infection while the remaining 20 cases (38.5%) were either of unknown etiology or caused by a combination of infectious agents.

Of the 46 patients with other organ specific and non-organ specific causes of death, 30 died from organ failure or dysfunction not caused by graft rejection, graft failure, cancer or infection, including complications to diabetes mellitus (N=9) (the complications include hypoglycemic episodes leading to death or universal arteriosclerosis leading to myocardial infarction and death), chronic obstructive pulmonary disease/pulmonary failure (N=8) and a variety of other specific causes. In 13 of the 30 recipients, the organ dysfunction leading to death was present prior to the transplantation. The remaining 16 recipients primarily died from hemorrhages, suicide, and alcohol abuse.

### Changes in specific causes of death over transplant year, transplant era and posttransplant period

Fig 2 demonstrates changes in the causes of death according to transplant era while changes by posttransplant period can be found in S2 Fig. Fig 3 shows adjusted hazard ratios for cause-specific death per one-year increase in transplant year.

**Fig 2.**
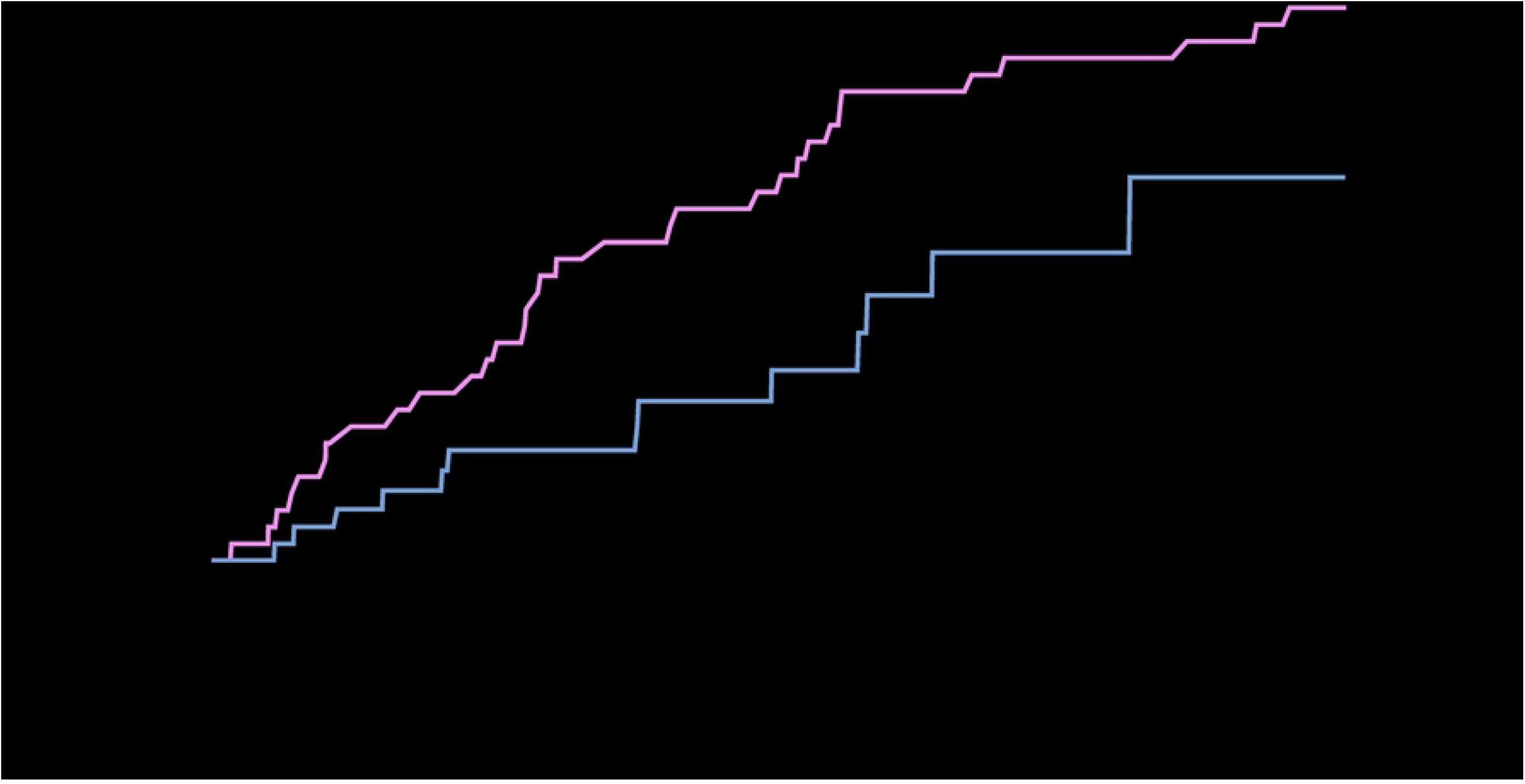
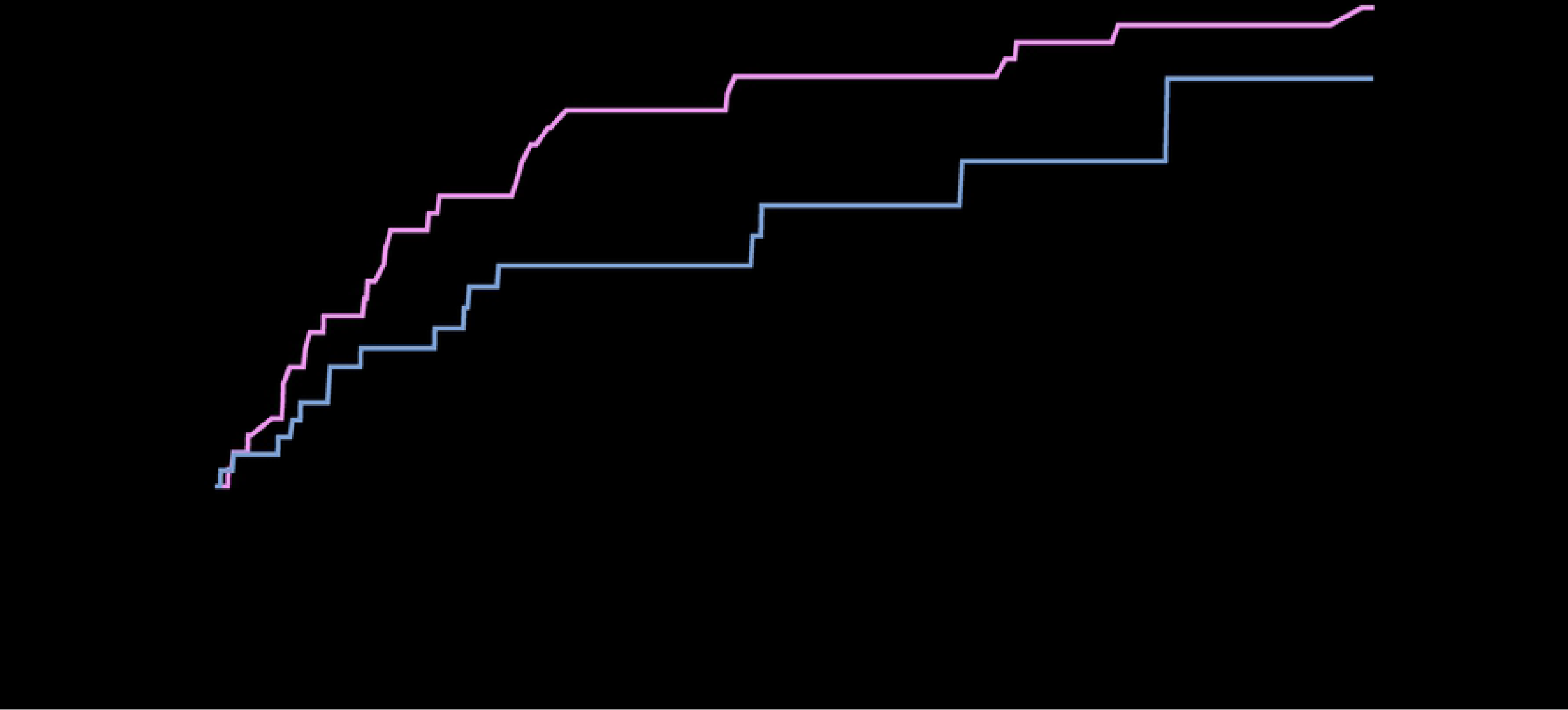
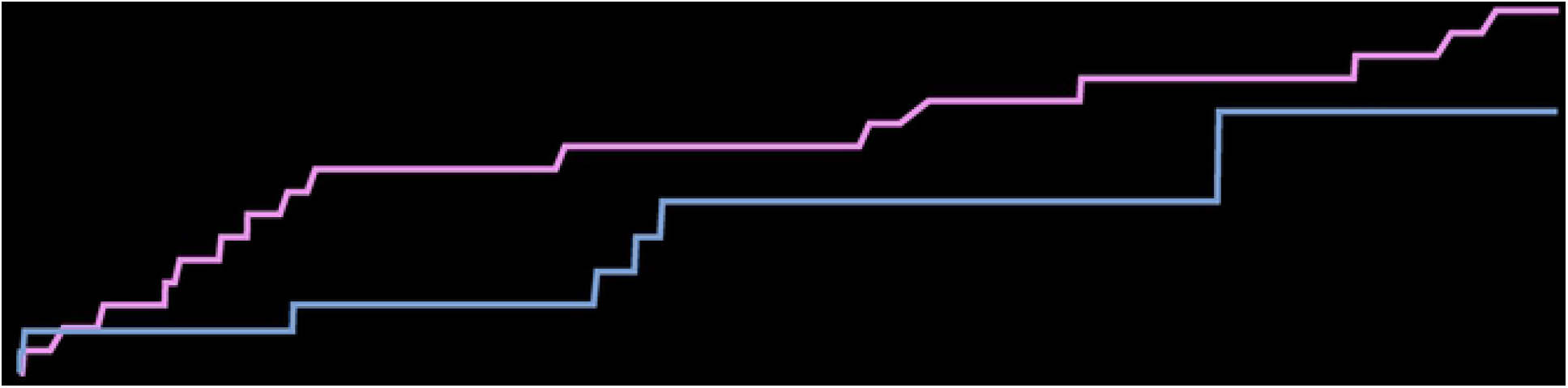
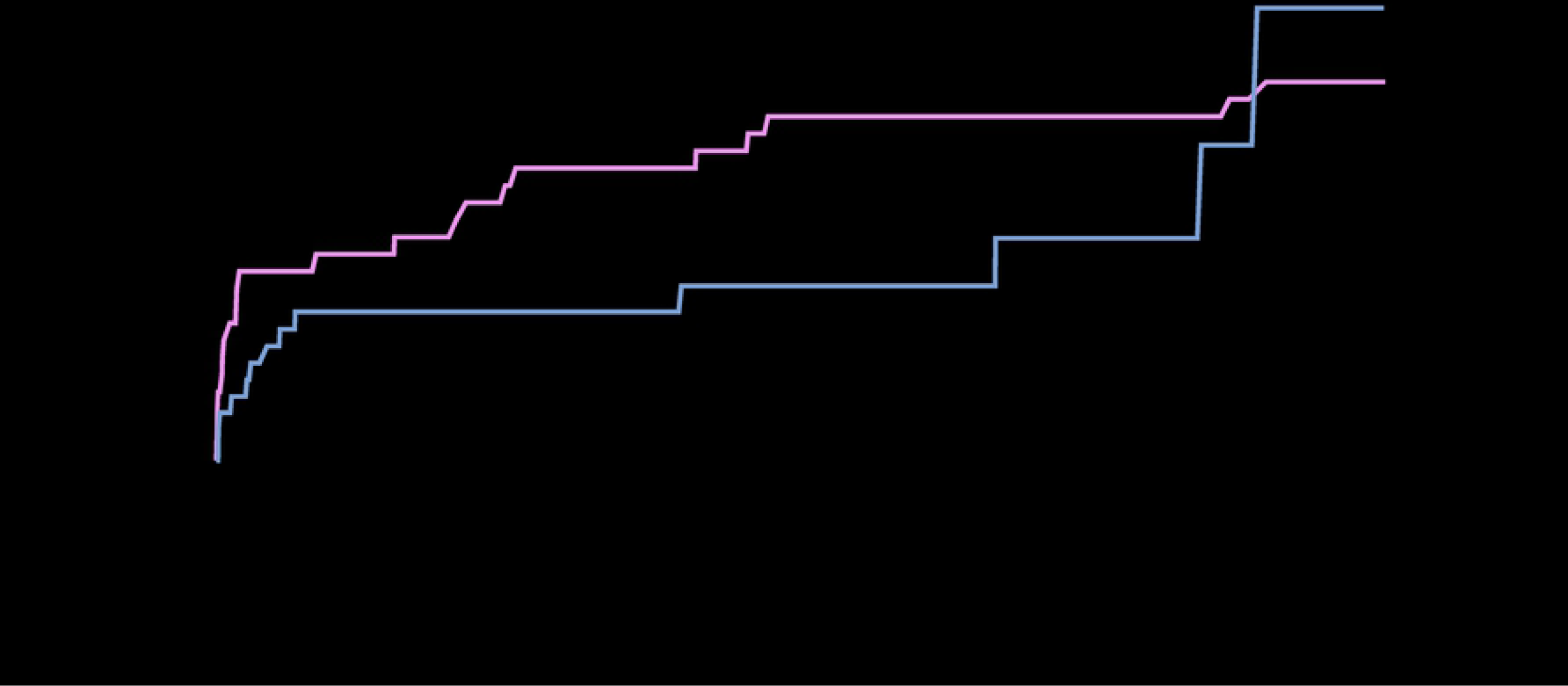
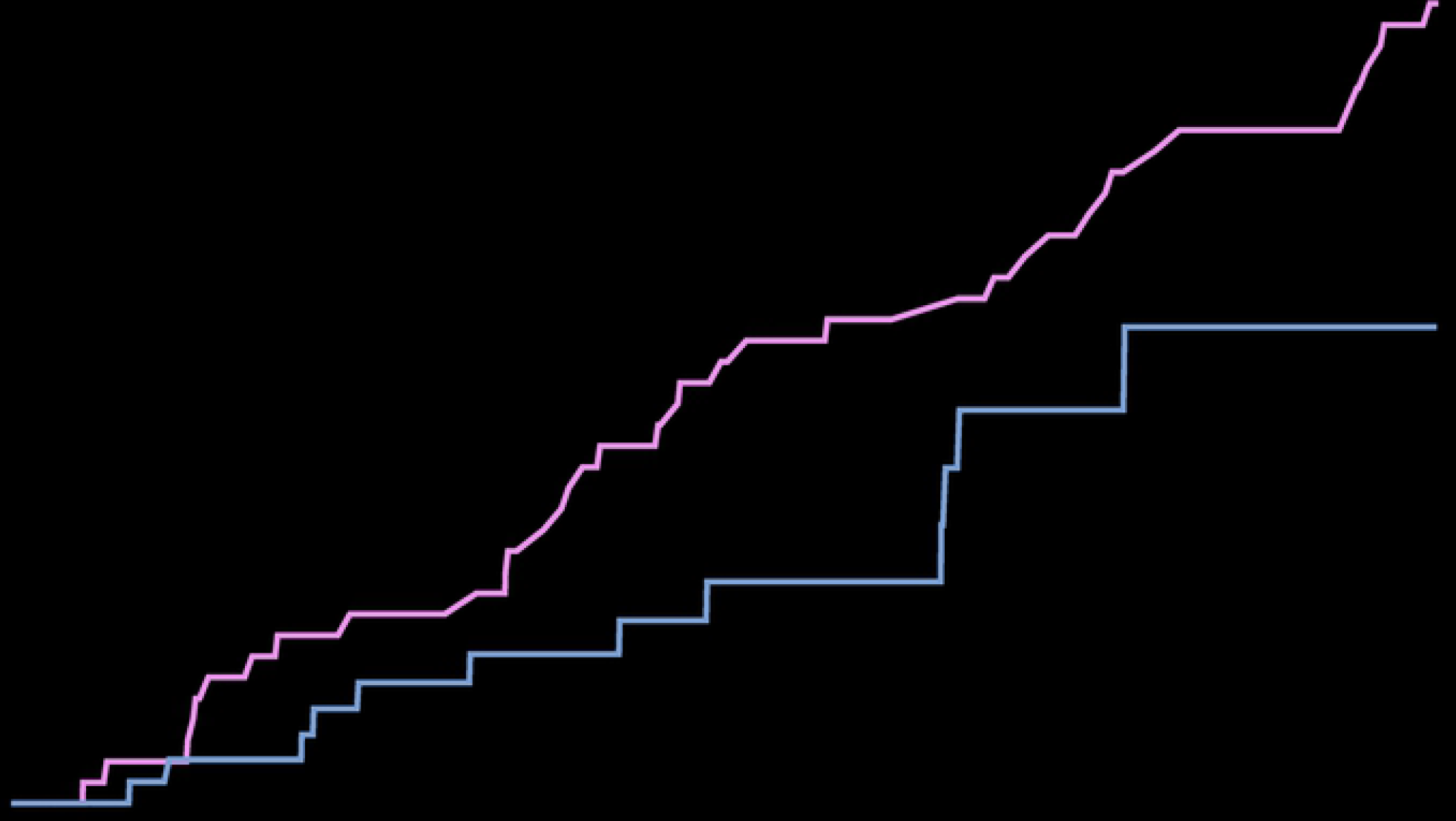
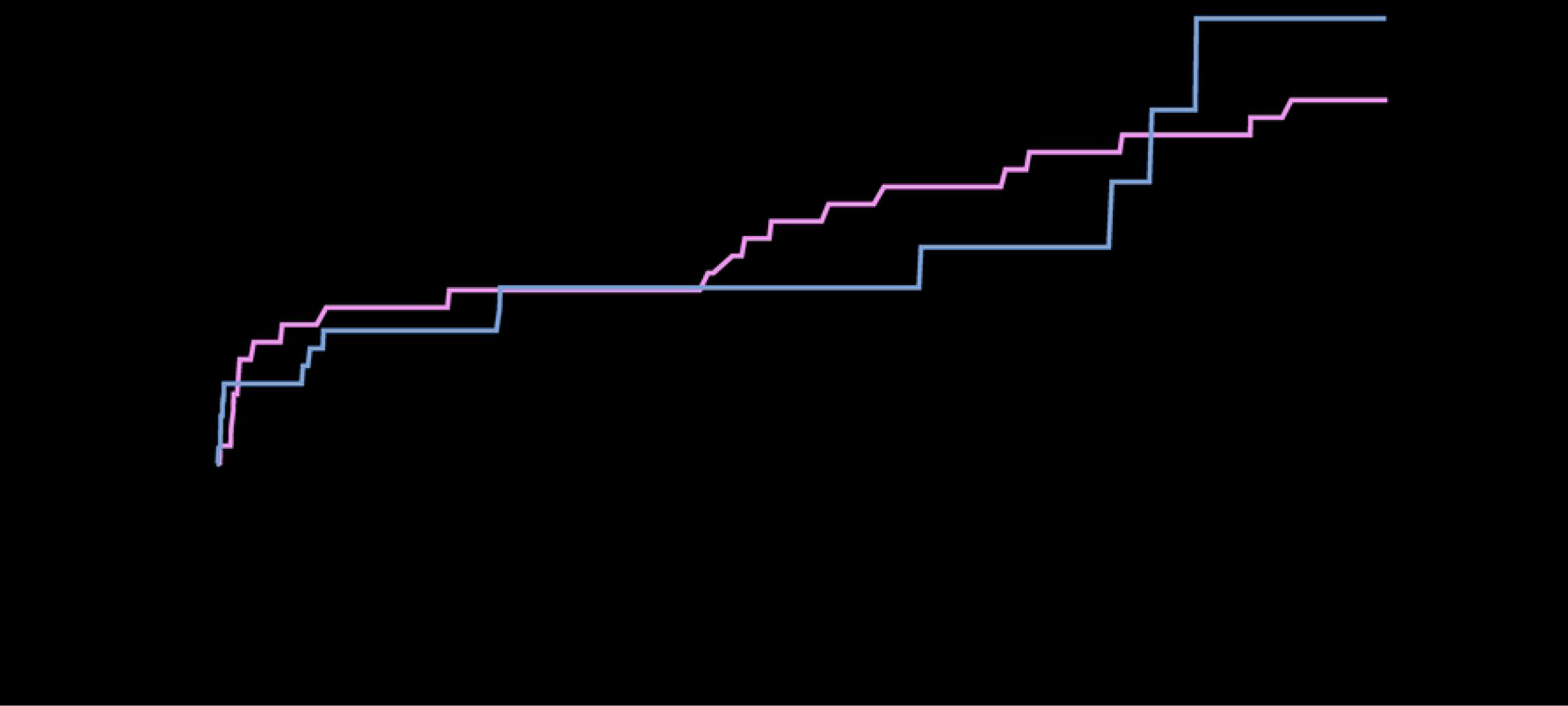
Cumulative incidence curves stratified by era of transplantation (2010-14 and 2015-19) for (A) death from graft rejection*, (B) death from infection, (C) death from cardiovascular disease, (D) death from graft failure, (E) death from cancer and (F) death from other organ specific and non-organ specific causes**. * Graft rejection includes one death from GvHD. ** Other organ specific and non-organ specific causes includes death from organ failure or dysfunction not caused by graft rejection, graft failure, cancer or infection, death from hemorrhage and death from other causes.

**Fig 3.**
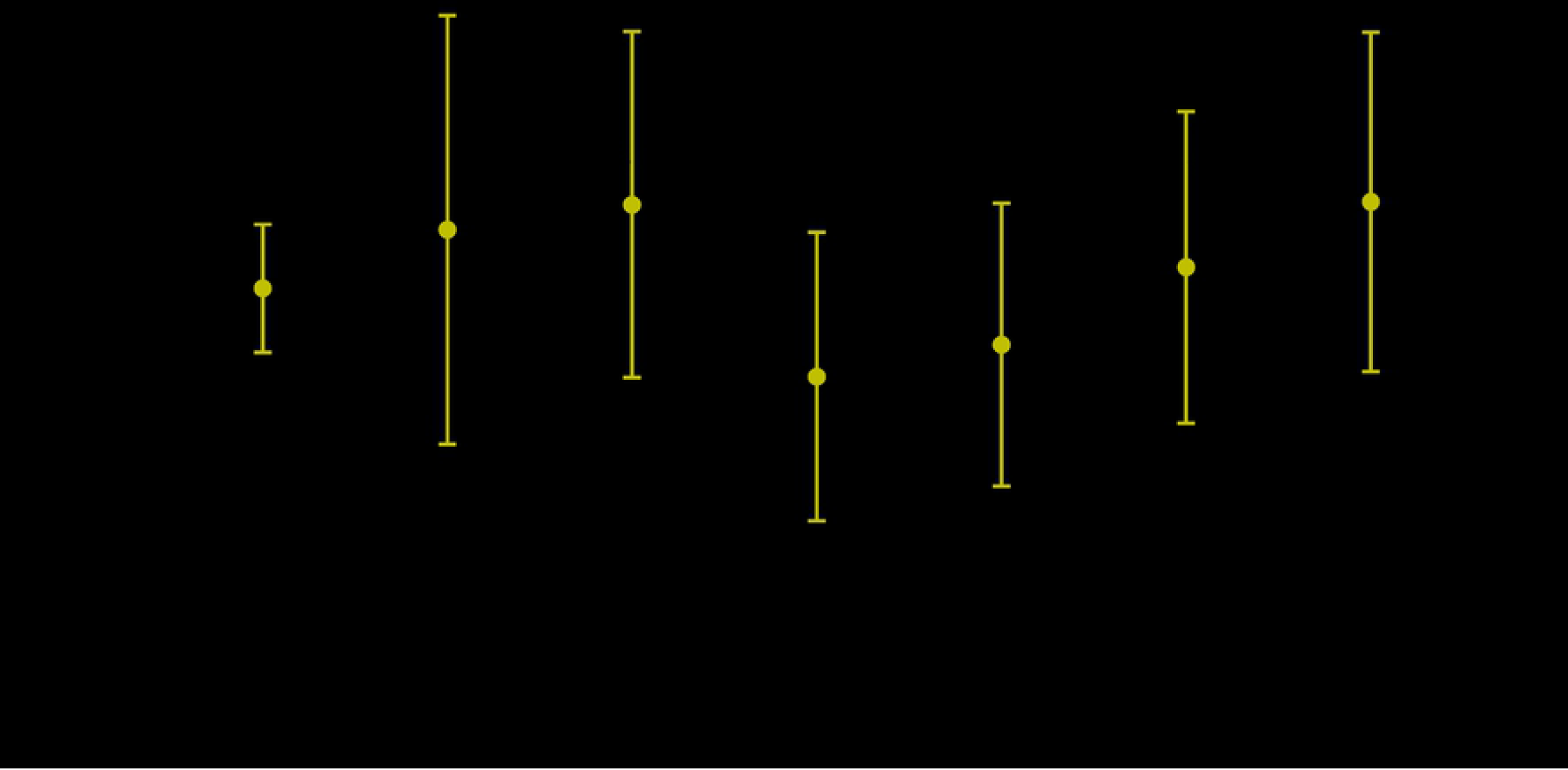
Adjusted hazard ratio* of mortality according to transplant calendar year (per one-year increase). * Adjusted for the baseline (gender, age at time of transplantation and transplanted organ) and time-updated characteristics (number of transplantations). ** Graft rejection includes one death from GvHD. *** Other organ specific and non-organ specific causes includes death from organ failure or dysfunction not caused by graft rejection, graft failure, cancer or infection, death from hemorrhage and death from other causes.

The cumulative incidence of graft rejection related death and infection related death decreased markedly from the early to the recent transplant era (Fig 2A-B). Thus, the cumulative incidence rate (CIR) of death from graft rejection at three years posttransplant was 3.3% (95% CI 2.2%-4.6%) in the early era and 1.9% (95% CI 1.0%-3.3%) in the recent era while the CIR of death from infections decreased from 2.8% (95% CI 1.9%-4.1%) to 1.9% (95% CI 1.1%-3.2%) over the two eras. Further, there was a trend towards lower cumulative incidence across transplant eras of death from cardiovascular disease, graft failure and cancer (Fig 2). Conversely, death from other organ specific and non-organ specific causes did not tend to decrease across the two transplant eras. Similar patterns were observed when transplanted organs were assessed separately, although numbers of deaths were too small to draw any conclusions.

There was a lower risk of graft rejection and infection related deaths by increasing transplant year after adjustment for baseline clinical characteristics (HR 0.84, 95% CI 0.74-0.95 and HR 0.86, 95% CI 0.77-0.97, respectively) (Fig 3). No other specific causes of death showed a statistically significant lower hazard of mortality over transplant year (Fig 3). A lower hazard of graft rejection related death with increasing transplant year was also observed in analyses including lung transplanted patients only (HR 0.83, 95% CI 0.72-0.95).

In the early posttransplant period (the first year after transplantation), the most common cause of death was infections (CIR 1.6%, 95% CI 1.0%-2.2% at 1 year posttransplant). In the following period, between >1 and 3 years after transplantation, graft rejection accounted for most deaths (CIR 1.9%, 95% CI 1.3%-2.8% at 3 years posttransplant). In the late period, between >3 and 10 years after transplantation, cancer (CIR 3.5%, 95% CI 2.3%-5.2%) and other organ specific and non-organ specific causes (CIR 3.3%, 95% CI 1.9%-5.4% at 10 years posttransplant) emerged as frequent causes of death (S2 Fig).

## Discussion

This study investigated trends in underlying cause of death in SOT recipients transplanted between 2010 and 2020 at a large tertiary transplant centre in Denmark, with close to complete ascertainment of death and 92% known cause of death. A lower risk of all-cause mortality was observed among individuals transplanted more recently, even after accounting for demographic characteristics and pre-transplant chronic diseases. This decrease was mainly driven by a decrease in deaths from graft rejection and death from infections, with death causes overall becoming increasingly diverse in more recent years.

The decreasing risk of graft rejection related deaths by transplant year was mainly driven by a decrease in this cause of death in lung transplant recipients. As most graft rejection related deaths were due to chronic graft rejection which often occur long after transplantation, it could be argued that the lower graft rejection related deaths in more recent transplant years was due to insufficient follow-up time. However, sensitivity analyses excluding patients transplanted in more recent years showed consistent results. Our observation is in line with previous reports showing a decrease in the incidence of chronic allograft rejection in lung transplant recipients in the past decade[4, 26].

We found a decline in risk of death from infections over the transplant years. Several recent large cohort studies have reported a similar decrease in infection related death after SOT in the past decade, most significantly in liver and kidney transplant recipients[27-29]. This likely reflects advances in immunosuppressive regimens and continuously increasing focus on primary prophylaxis and screening for emerging infection before causing serious disease, allowing for pre-emptive therapy.

While risk of death due to infection and graft rejection decreased significantly over the transplant years, there was a relative increase in the diversity of the causes of death in the present study. Thus, deaths due to a wide range of organ failure or dysfunction not caused by cancer, graft failure, rejection, or infections was the only category of death causes in this study that did not show a decreasing tendency between the early and more recent transplant eras. The increase in cause-of-death variation is likely due to a decrease in risk of death from infection and graft rejection which leads to a parallel relative increase in diverse causes of death. Moreover, as both surgical and medical treatment improves, transplantation indications have expanded in recent years and transplant candidates more often have pre-transplant comorbidities[30]. Thus, recipients may be more prone to developing various end-organ diseases in more recent years suggesting a higher competing risk between diseases to be registered as the cause of death. In fact, almost half of the recipients of our study who died from end-organ failure not caused by cancer, graft failure, rejection or infections suffered from these chronic diseases at time of transplantation. Notably, as opposed to what has been reported in previous studies, we observed no increase in pre-transplant comorbidities over time in our cohort. This may reflect the unavailability of comorbidity data after 2017 and could thus be due to lack of power. Further studies are warranted to better understand the relationship between chronic diseases at time of transplantation and cause of death.

Although the cumulative incidence of death from cancer seemed to decrease in the more recent transplant era, the risk of cancer death remained relatively stable over time after adjusting for potential confounders. Most previous studies have reported an increase in cancer death over calendar periods which is likely a function of the long-term immunosuppression in long-term survivors and an increasingly ageing transplant population[31, 32]. The lack of long-term follow-up in patients transplanted in the more recent years in the present study could explain the lack of confirmation of previous observations.

The underlying cause of death could be determined in the majority of the patients and only 8% in this study were classified with an unknown cause of death due to insufficient information. In comparison, deaths classified as unknown or missing cause have been reported in up to 40% of early deaths and 61.5% of late deaths in previous studies[13, 16-19] potentially overlooking important tendencies.

A major strength of this study is the reliable classification of specific, underlying causes of death using the validated CLASS method[20]. Using consistent reviewers and CLASS-investigators across time periods, the method has been utilized in the classification of every death in this study. The method provides a uniform classification system and involves systematic review of medical files and assessment by transplant specialists. Another strength is that we included a large cohort of almost unselected patients at a large regional transplant center.

The present study has some limitations. The study includes a heterogeneous group of transplanted patients over a long time with differing underlying risk factors and transplant indications. Thus, we were generally unable to assess trends in cause of death in specific transplanted organs. Further, we were unable to adjust for changes in immunosuppressive treatment regimens which may impact death from especially infections and graft rejection. The length of the study period renders it difficult to assess death causes that may occur 5-10 years post transplantation. Thus, changes over time in deaths from cancer and other causes which may take longer time to manifest themselves should be interpreted with caution. As is the case with observational studies, there may be other relevant factors such as additional pretransplant comorbidities or yet unidentified confounders which could not be considered in these analyses and may also have an impact on the results presented.

In conclusion, the study shows a significant decrease in risk of all-cause mortality for solid organ transplant recipients over transplant year between 2010 and 2020, which was mainly driven by a decreasing risk of death from graft rejection and infections. On the other hand, the relative variation and diversity of cause of death showed a tendency to increase over the transplant years. In many cases the chronic disease leading to death was present at time of transplantation. Increasing variation in cause of death is potentially an important concern as the cause of mortality becomes more unpredictable. Greater variation in causes of death requires additional focus on establishing a clinical infrastructure to monitor a wider range of morbidity and mortality causes in the coming years.

## Data Availability

The authors cannot disclose individual level data because of ethical and legal data protection restrictions and have included group level data in the paper and tables. The dataset from this study is held securely in coded form at Centre of Excellence for Health, Immunity and Infections (CHIP). While national data protection laws and ethical regulations as well as cooperation agreements prohibit CHIP from making the dataset publicly available, requests for access to anonymized or pooled data may be sent to the Data Access Committee of CHIP.

## Acknowledgement

We would like to thank Jacob Dam Svendsen, Torben Møller Weide and Erik Viuff Hansen for excellent data management assistance, and the MATCH steering committee for providing access to the MATCH database.

## Supporting information

**S1 Table. Data sources and dates**.

**S1 Fig. Association between baseline characteristics and all-cause mortality**.

*Results are shown on a log(2)-scale. **Per one year increase in transplant year.

**S2 Fig. Cumulative incidence curves for specific causes of death* according to posttransplant time periods. (A) ≤1 year posttransplant, (B) between >1 and 3 years posttransplant, (C) between >3 and 10 years posttransplant**. *Graft rejection includes one death from GvHD. Other organ specific and non-organ specific causes includes death from organ failure or dysfunction not caused by graft rejection, graft failure, cancer or infection, death from hemorrhage and death from other causes.

## References

1. Adam R, Karam V, Cailliez V, Jg OG, Mirza D, Cherqui D, et al. 2018 Annual Report of the European Liver Transplant Registry (ELTR) - 50-year evolution of liver transplantation. Transpl Int. 2018;31(12):1293–317. Epub 2018/09/28. doi: 10.1111/tri.13358. PubMed PMID: 30259574.

2. Lund LH, Khush KK, Cherikh WS, Goldfarb S, Kucheryavaya AY, Levvey BJ, et al. The Registry of the International Society for Heart and Lung Transplantation: Thirty-fourth Adult Heart Transplantation Report-2017; Focus Theme: Allograft ischemic time. J Heart Lung Transplant. 2017;36(10):1037–46. Epub 2017/08/07. doi: 10.1016/j.healun.2017.07.019. PubMed PMID: 28779893.

3. Briggs JD. Causes of death after renal transplantation. Nephrol Dial Transplant. 2001;16(8):1545–9. Epub 2001/07/31. doi: 10.1093/ndt/16.8.1545. PubMed PMID: 11477152.

4. Chambers DC, Cherikh WS, Harhay MO, Hayes D, Jr., Hsich E, Khush KK, et al. The International Thoracic Organ Transplant Registry of the International Society for Heart and Lung Transplantation: Thirty-sixth adult lung and heart-lung transplantation Report-2019; Focus theme: Donor and recipient size match. J Heart Lung Transplant. 2019;38(10):1042–55. Epub 2019/09/25. doi: 10.1016/j.healun.2019.08.001. PubMed PMID: 31548030; PubMed Central PMCID: PMCPMC6816340.

5. Atabani SF, Smith C, Atkinson C, Aldridge RW, Rodriguez-Perálvarez M, Rolando N, et al. Cytomegalovirus replication kinetics in solid organ transplant recipients managed by preemptive therapy. Am J Transplant. 2012;12(9):2457–64. Epub 2012/05/19. doi: 10.1111/j.1600-6143.2012.04087.x. PubMed PMID: 22594993; PubMed Central PMCID: PMCPMC3510308.

6. Kotton CN. CMV: Prevention, Diagnosis and Therapy. Am J Transplant. 2013;13 Suppl 3:24–40; quiz Epub 2013/02/01. doi: 10.1111/ajt.12006. PubMed PMID: 23347212.

7. Sato K, Ogawa K, Onumata O, Aso K, Nakayama Y, Yoshida K, et al. Cause of death in renal transplant patients: a comparison between azathioprine and ciclosporin. Surg Today. 2001;31(8):681–7. Epub 2001/08/21. doi: 10.1007/s005950170070. PubMed PMID: 11510603.

8. Barber K, Blackwell J, Collett D, Neuberger J. Life expectancy of adult liver allograft recipients in the UK. Gut. 2007;56(2):279–82. Epub 2006/09/30. doi: 10.1136/gut.2006.093195. PubMed PMID: 17008365; PubMed Central PMCID: PMCPMC1856771.

9. Gelson W, Hoare M, Dawwas MF, Vowler S, Gibbs P, Alexander G. The pattern of late mortality in liver transplant recipients in the United Kingdom. Transplantation. 2011;91(11):1240–4. Epub 2011/04/26. doi: 10.1097/TP.0b013e31821841ba. PubMed PMID: 21516069.

10. Durand F, Levitsky J, Cauchy F, Gilgenkrantz H, Soubrane O, Francoz C. Age and liver transplantation. J Hepatol. 2019;70(4):745–58. Epub 2018/12/24. doi: 10.1016/j.jhep.2018.12.009. PubMed PMID: 30576701.

11. McCurry KR. Brief Overview of Lung, Heart, and Heart-Lung Transplantation. Crit Care Clin. 2019;35(1):1–9. Epub 2018/11/19. doi: 10.1016/j.ccc.2018.08.005. PubMed PMID: 30447772.

12. Recipients SRoT. OPTN/SRTR 2019 Annual Data Report: Kidney Scientific Registry of Transplant Recipients 2021 [cited 2022 January 10th]. Available from: https://www.srtr.org/reports/srtroptn-annual-data-report/.

13. Awan AA, Niu J, Pan JS, Erickson KF, Mandayam S, Winkelmayer WC, et al. Trends in the Causes of Death among Kidney Transplant Recipients in the United States (1996-2014). Am J Nephrol. 2018;48(6):472–81. Epub 2018/11/26. doi: 10.1159/000495081. PubMed PMID: 30472701; PubMed Central PMCID: PMCPMC6347016.

14. Kirklin JK, Naftel DC, Bourge RC, McGiffin DC, Hill JA, Rodeheffer RJ, et al. Evolving trends in risk profiles and causes of death after heart transplantation: a ten-year multi-institutional study. J Thorac Cardiovasc Surg. 2003;125(4):881–90. Epub 2003/04/17. doi: 10.1067/mtc.2003.168. PubMed PMID: 12698152.

15. Na R, Grulich AE, Meagher NS, McCaughan GW, Keogh AM, Vajdic CM. De novo cancer-related death in Australian liver and cardiothoracic transplant recipients. Am J Transplant. 2013;13(5):1296–304. Epub 2013/03/08. doi: 10.1111/ajt.12192. PubMed PMID: 23464511.

16. UK Renal Registry (2020) UK Renal Registry Summary of Annual Report – analyses of adult data to the end of 2018, Bristol, UK.. United Kingdom Kidney Association: United Kingdom Kidney Association Registry UR; 2020.

17. Acuna SA, Fernandes KA, Daly C, Hicks LK, Sutradhar R, Kim SJ, et al. Cancer Mortality Among Recipients of Solid-Organ Transplantation in Ontario, Canada. JAMA Oncol. 2016;2(4):463–9. Epub 2016/01/10. doi: 10.1001/jamaoncol.2015.5137. PubMed PMID: 26746479.

18. El-Husseini A, Aghil A, Ramirez J, Sawaya B, Rajagopalan N, Baz M, et al. Outcome of kidney transplant in primary, repeat, and kidney-after-nonrenal solid-organ transplantation: 15-year analysis of recent UNOS database. Clin Transplant. 2017;31(11). Epub 2017/09/08. doi: 10.1111/ctr.13108. PubMed PMID: 28881060.

19. Punnoose LR, Rao S, Ghanta MM, Karhadkar SS, Alvarez R. Outcomes of Older Patients in the Recent Era of Heart Kidney Transplantation. Transplant Proc. 2021;53(1):341–7. Epub 2020/07/23. doi: 10.1016/j.transproceed.2020.04.1821. PubMed PMID: 32694056.

20. Wareham NE, Da Cunha-Bang C, Borges Á H, Ekenberg C, Gerstoft J, Gustafsson F, et al. Classification of death causes after transplantation (CLASS): Evaluation of methodology and initial results. Medicine (Baltimore). 2018;97(29):e11564. Epub 2018/07/20. doi: 10.1097/md.0000000000011564. PubMed PMID: 30024557; PubMed Central PMCID: PMCPMC6086480.

21. Ekenberg C, da Cunha-Bang C, Lodding IP, Sørensen SS, Sengeløv H, Perch M, et al. Evaluation of an electronic, patient-focused management system aimed at preventing cytomegalovirus disease following solid organ transplantation. Transpl Infect Dis. 2020;22(2):e13252. Epub 2020/01/31. doi: 10.1111/tid.13252. PubMed PMID: 31997565.

22. MATCH (Management of Post-Transplant Infections in Collaborating Hospitals): Rigshospitalet, University of Copenhagen Centre of Excellence for Health, Immunity and Infections (CHIP), Section 2100; [cited 2022 January 10th]. Available from: https://chip.dk/Clinical-programs/MATCH.

23. Schmidt M, Pedersen L, Sorensen HT. The Danish Civil Registration System as a tool in epidemiology. Eur J Epidemiol. 2014;29(8):541–9. Epub 2014/06/27. doi: 10.1007/s10654-014-9930-3. PubMed PMID: 24965263.

24. Cause of Death List: Rigshospitalet, University of Copenhagen Centre of Excellence for Health, Immunity and Infections (CHIP); 2021 [cited 2022 January 10th]. Available from: https://chip.dk/Portals/0/CLASS_Cause%20of%20Death%20List%202021.pdf?ver=2021-10-13-105332-330&timestamp=1634115217608.

25. Centre of Excellence for Personalised Medicine of Infectious Complications in Immune Deficiency (PERSIMUNE): Rigshospitalet, University of Copenhagen Centre of Excellence for Health, Immunity and Infections (CHIP); [cited 2022 January 10th]. Available from: http://www.persimune.dk/.

26. Costa J, Benvenuto LJ, Sonett JR. Long-term outcomes and management of lung transplant recipients. Best Pract Res Clin Anaesthesiol. 2017;31(2):285–97. Epub 2017/11/08. doi: 10.1016/j.bpa.2017.05.006. PubMed PMID: 29110800.

27. Kinnunen S, Karhapää P, Juutilainen A, Finne P, Helanterä I. Secular Trends in Infection-Related Mortality after Kidney Transplantation. Clin J Am Soc Nephrol. 2018;13(5):755–62. Epub 2018/04/07. doi: 10.2215/cjn.11511017. PubMed PMID: 29622669; PubMed Central PMCID: PMCPMC5969482.

28. Rana A, Ackah RL, Webb GJ, Halazun KJ, Vierling JM, Liu H, et al. No Gains in Long-term Survival After Liver Transplantation Over the Past Three Decades. Ann Surg. 2019;269(1):20–7. Epub 2018/01/06. doi: 10.1097/sla.0000000000002650. PubMed PMID: 29303806.

29. Ying T, Shi B, Kelly PJ, Pilmore H, Clayton PA, Chadban SJ. Death after Kidney Transplantation: An Analysis by Era and Time Post-Transplant. J Am Soc Nephrol. 2020;31(12):2887–99. Epub 2020/09/11. doi: 10.1681/asn.2020050566. PubMed PMID: 32908001; PubMed Central PMCID: PMCPMC7790214.

30. Lund LH, Edwards LB, Kucheryavaya AY, Benden C, Dipchand AI, Goldfarb S, et al. The Registry of the International Society for Heart and Lung Transplantation: Thirty-second Official Adult Heart Transplantation Report--2015; Focus Theme: Early Graft Failure. J Heart Lung Transplant. 2015;34(10):1244–54. Epub 2015/10/12. doi: 10.1016/j.healun.2015.08.003. PubMed PMID: 26454738.

31. Acuna SA. Etiology of increased cancer incidence after solid organ transplantation. Transplant Rev (Orlando). 2018;32(4):218–24. Epub 2018/07/19. doi: 10.1016/j.trre.2018.07.001. PubMed PMID: 30017342.

32. Wimmer CD, Rentsch M, Crispin A, Illner WD, Arbogast H, Graeb C, et al. The janus face of immunosuppression - de novo malignancy after renal transplantation: the experience of the Transplantation Center Munich. Kidney Int. 2007;71(12):1271–8. Epub 2007/03/03. doi: 10.1038/sj.ki.5002154. PubMed PMID: 17332737.

